# Early Clinical Factors Predicting the Development of Critical Disease in Japanese Patients with COVID-19: A Single-Center Retrospective, Observational Study

**DOI:** 10.1101/2020.07.29.20159442

**Authors:** Takatoshi Higuchi, Tsutomu Nishida, Hiromi Iwahashi, Osamu Morimura, Yasushi Otani, Yukiyoshi Okauchi, Masaru Yokoe, Norihiro Suzuki, Masami Inada, Kinya Abe

## Abstract

**Background:** Insufficient evidence of factors predicting the COVID-19 progression from mild to moderate to critical has been established. We retrospectively evaluated risk factors for critical progression in Japanese COVID-19 patients.

**Method:** Seventy-four laboratory-confirmed COVID-19 patients were hospitalized in our hospital between February 20, 2020, and June 10, 2020. We excluded asymptomatic, non-Japanese, and child patients. We divided patients into the stable group (SG) and the progression group (PG) (patients requiring mechanical ventilation). We compared the clinical factors in both groups. We established the cutoff values (COVs) for significantly different factors via receiver operating characteristic (ROC) curve analysis and evaluated risk factors by univariate regression.

**Results:** We enrolled 57 COVID-19 patients (median age 52 years, 56.1% male). The median progression time from symptom onset was eight days. Seven patients developed critical disease (PG: 12.2%), two (3.5%) of whom died; 50 had stable disease. Univariate logistic analysis identified elevated lactate dehydrogenase (LDH) (COV: 309 U/l), decreased estimated glomerular filtration rate (eGFR) (COV: 68 ml/min), lymphocytopenia (COV: 980/μl), and statin use as significantly associated with disease progression. However, in Cox proportional hazards analysis, lymphocytopenia at symptom onset was not significant.

**Conclusions:** We identified three candidate risk factors for adult Japanese patients with mild to moderate COVID-19: statin use, elevated LDH level, and decreased eGFR.

## Introduction

In December 2019, a novel coronavirus outbreak occurred in Wuhan, China. In February 2020, the World Health Organization (WHO) named this disease, caused by severe acute respiratory syndrome coronavirus 2 (SARS-CoV-2), coronavirus disease 2019 (COVID-19). It has rapidly spread and resulted in a pandemic. The clinical presentation of COVID-19 patients ranges from asymptomatic to mild to critical, and the disease can lead to death. The WHO classified COVID-19 into four severity levels: mild, moderate, severe, and critical ^1^. The proportion of patients with severe or critical diseases may vary with location, but most patients have mild to moderate disease ^2,3^. However, some patients rapidly progress to critical illness ^4^. The mortality risk dramatically increases once a patient develops critical disease ^5^. Therefore, early prediction of critical COVID-19 development is required for timely intervention to improve prognosis. In addition, ethnic differences may affect the risk factors because of the different extent of the pandemic. However, there is no established evidence of predictive factors for developing critical COVID-19 in the Japanese population. Our hospital is a medium-volume hospital with 613 beds, including 14 in the Infectious Disease Unit, in an urban area of Osaka Prefecture, Japan. It is also designated as a medical institution for type II infectious diseases, of which there are 351 with a total of 1758 beds in Japan. We have accepted COVID-19 patients since February 2020 and have expanded the number of beds for infectious disease patients from 14 to 45 to allow more COVID-19 patients. In the present study, we retrospectively examined the risk factors for progression to a critical stage in Japanese patients with COVID-19 using clinical characteristics and laboratory findings in a single hospital.

## Patients and methods

This study was a retrospective single-center cohort study that examined a total of 75 consecutive patients with laboratory-confirmed COVID-19 from February 20, 2020, until June 10, 2020. We followed the patients until July 7, 2020. In the present study, various reverse transcription-polymerase chain reaction (RT-PCR) assays were used, but all findings were based on the results of tests by public health centers in the Osaka area in Japan. Osaka Prefecture established the Osaka Prefecture Inpatient Follow-up Center to coordinate broad-based hospitalization based on patients’ symptoms on March 30, 2020. Patients with confirmed or suspected COVID-19 were triaged in four stages depending on the disease severity: 1) patients with critical illness were hospitalized at designated medical institutions for infectious diseases, university hospitals, and the National Hospital Organization; 2) patients with moderate disease were hospitalized in a general hospital (negative pressure room or special ward); 3) patients with mild disease were isolated in a closed or decommissioned ward; and 4) asymptomatic pathogen carriers were followed up in private accommodations or at home. Our hospital was designated a general hospital to treat moderate COVID-19 patients.

We reviewed medical records and examined the clinical factors that predict progression to critical disease among patients with mild to moderate COVID-19. According to the clinical course, we divided the patients into two groups: the stable group (SG) and the progression group (PG).

The present study was conducted in accordance with the Declaration of Helsinki, and approval was obtained from the Institutional Review Board of Toyonaka Municipal Hospital (No. 2020-07-01). The requirement for informed consent was waived via the *opt-out* method on our hospital website.

### Medical treatment

We provided basic supportive care, including supplemental oxygen, and the use of acetaminophen, antibiotics, and mechanical ventilatory support when indicated. Regarding specific medications, we used ciclesonide for patients with lung infiltrates on imaging, hydroxychloroquine for patients with a high fever (higher than 38°C) or diarrhea, and favipiravir for patients with moderate pneumonia, according to the advice of respiratory disease specialists.

### Definition

A diagnosis of laboratory-confirmed COVID-19 was made in cases of positive direct detection of SARS-CoV-2 nucleic acid by RT-PCR ^6^. The severity of COVID-19 was defined as follows: 1) mild to moderate disease with or without pneumonia; 2) severe disease with dyspnea, a respiratory frequency of ≥ 30 breaths/minute, blood oxygen saturation of ≤ 93%, PaO2/FiO2 ratio of <300, and/or lung infiltrates in >50% of the lung field within 24-48 hours; or 3) critical disease with respiratory failure, septic shock, or multiple organ dysfunction/failure according to the criteria in the Report of the WHO-China Joint Mission on COVID-19 ^1^. In the present study, progression was defined when a patient required mechanical ventilatory support.

### Statistical analysis

The medians and interquartile ranges (IQRs) are reported for continuous variables. Categorical variables are summarized as frequencies (percentages). Missing continuous variables were imputed using a matrix imputation method, and missing categorical variables were imputed using random proportional methods. Differences in variables between the SG and PG were evaluated using Fisher’s exact test for categorical data and the Mann-Whitney U test for continuous data. To determine the significant factors for differentiating between the SG and PG, we used receiver operating characteristic (ROC) curve analysis to determine the cutoff values (COVs) for predicting disease progression. Then, we used a univariate logistic regression model to obtain the odds ratios (ORs) with 95% confidence intervals (CIs) between the two groups according to the COVs for the significant factors. We defined the observation time as the interval from the onset of symptoms to the development of critical disease or discharge. The cumulative rates of progression to critical disease were calculated using the Kaplan–Meier method and compared using the log-rank test. Similarly, we used univariate Cox proportional hazards models to obtain the hazard ratios (HRs) with 95% CIs.

All reported P values were two-sided, and P<0.05 was considered significant. Statistical analyses were performed with JMP statistical software (ver. 14.3, SAS Institute, Inc., Cary, NC, USA).

## Results

### Baseline characteristics and clinical course

All consecutive COVID-19 patients hospitalized at Toyonaka Municipal Hospital between February 20 and June 10, 2020, were enrolled. There were a total of 74 patients with laboratory-confirmed COVID-19. Among these patients, we excluded seven asymptomatic patients, five readmitted patients, one patient transferred from another hospital, seven non-Japanese patients, and three patients younger than 16 years of age. Figure 1 shows a flow chart of patient enrollment. By the end of the follow-up period, all patients had been discharged, and we ultimately enrolled a total of 57 patients with newly diagnosed, laboratory-confirmed, mild to moderate or severe COVID-19.

**Figure 1.**
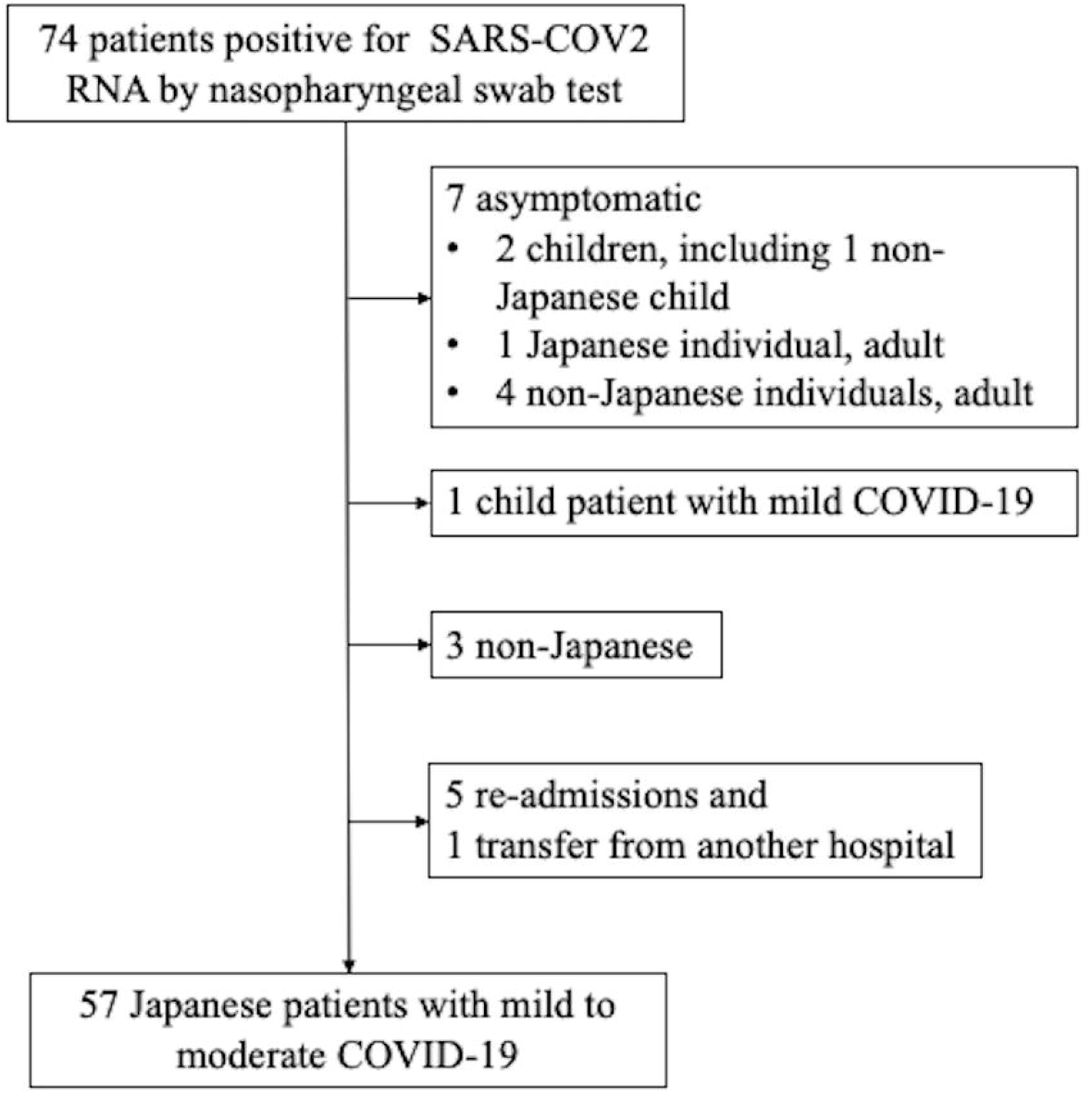
Flow chart of patient enrollment.

Table 1 shows the patients’ comorbidities, medication use, and treatment and clinical course during hospitalization. Of the 57 patients with mild to moderate COVID-19, 32 (56.1%) were male, and the median (interquartile range (IQR)) age was 52 (35, 69.5) years. Sixteen patients (28.1%) had a history of close contact with individuals with confirmed COVID-19 cases. At admission, the median disease duration from symptom onset was 8 (5, 12) days.

**Table 1.** Characteristics and clinical course of patients with mild to moderate confirmed COVID-19

| Characteristics | Patients with mild to moderate COVID-19<br>N=57 |
| --- | --- |
| Age, median (IQR) | 52 (35, 69.5) |
| Male Sex, n (%) | 32 (56.1) |
| Body mass index, median (IQR) | 23.8 (21.0, 26.5) |
| Pneumonia, n (%) | 37 (64.9) |
| Smoking history (Non/past/current) | 33/17/7 |
| History of close contact with individuals with confirmed cases | 16 (28.1) |
| Days from onset of symptoms to admission, median (IQR) | 8 (5, 12) |
| Comorbidities |  |
| Hypertension, n (%) | 16 (28.1) |
| Cardiovascular diseases, n (%) | 5 (8.8) |
| Chronic obstructive pulmonary disease, n (%) | 4 (7.0) |
| Asthma, n (%) | 8 (14.0) |
| Diabetes mellitus, n (%) | 13 (22.8) |
| Hyperlipidemia, n (%) | 20 (35.1) |
| Chronic kidney disease, n (%) | 5 (8.8) |
| Hemodialysis, n (%) | 3 (5.3) |
| Solid cancer, n (%) | 1 (1.8) |
| Pregnancy, n (%) | 2 (3.5) |
| Use of medication for comorbidities |  |
| ARB, n (%) | 8 (14.0) |
| Calcium blocker, n (%) | 9 (15.8) |
| Statin, n (%) | 12 (21.1) |

Most patients (89.9%) had a fever at disease onset; the second most common symptom was diarrhea (25.9%). A total of 37 (64.9%) and 20 (35.1%) patients had mild to moderate disease and severe disease, respectively. Regarding the medications used to treat COVID-19, we treated 29 patients with ciclesonide, 14 with hydroxychloroquine, and 12 with favipiravir (Table 2).

**Table 2.** Initial presentation, treatment, and clinical course

| Initial presentation |  |
| --- | --- |
| Fever, n (%) | 51 (89.9) |
| Fatigue, n (%) | 12 (21.1) |
| Cough, n (%) | 5 (8.8) |
| Dyspnea, n (%) | 13 (22.8) |
| Sputum production, n (%) | 5 (8.8) |
| Anorexia, n (%) | 7 (12.3) |
| Headache, n (%) | 5 (8.8) |
| Diarrhea, n (%) | 14 (25.9) |
| New loss of taste or smell, n (%) | 9 (15.8) |
| Erythema, n (%) | 3 (5.3) |
| Severity of COVID-19 |  |
| Mild to moderate/Severe, n (%) | 37 (64.9)/20 (35.1) |
| Treatment |  |
| Required oxygen, n (%) | 20 (35.1) |
| Medication for COVID-19 |  |
| Ciclesonide, n (%) | 29 (50.9) |
| Hydroxychloroquine, n (%) | 14 (24.6) |
| Favipiravir, n (%) | 12 (21.1) |
| Clinical course |  |
| Length of hospital stay, median (IQR) (days) | 12 (8, 20) |
| Required mechanical ventilatory support, n (%) | 7 (12.3) |
| Mortality, n (%) | 2 (3.5) |

The median hospital stay was 12 (8, 20) days. During the hospital stay, seven patients developed critical COVID-19 (PG: 12.2%), and 50 patients did not experience progression to critical status (SG: 78.8%). The median time from symptom onset to disease progression was nine days, and the time from admission to progression was one day. Most cases rapidly progressed to critical status after admission, but one 61-year-old woman developed critical illness 20 days after the onset of symptoms and nine days after admission. She had a case of hospital-acquired SARS-CoV-2 infection in a previous hospital while she was being treated with high-dose steroid therapy for nephrotic syndrome. In the PG group, two patients (3.5%) died of COVID-19. Both were men aged 70 years or older (71 and 75 years old) with diabetes.

### Risk factors affecting disease progression

The PG had significantly higher statin use, a lower lymphocyte count, a higher platelet count, a higher lactate dehydrogenase (LDH) level, a higher C-reactive protein (CRP) level, a higher aspartate aminotransferase (AST) level, a lower creatinine (Cr) level, a higher blood urea nitrogen (BUN) level, and a lower estimated glomerular filtration rate (eGFR) than the SG. We evaluated the COVs of these laboratory data for predicting disease progression via ROC curve analysis. Our analysis demonstrated that the COVs for the lymphocyte count, LDH level, CRP level, and eGFR were 980 (area under the ROC curve (AUC): 0.85, sensitivity=1.00, and specificity=0.62), 309 (AUC: 0.81, sensitivity=0.857, and specificity=0.70), 2.92 mg/dl (AUC: 0.76, sensitivity=1.00, and specificity=0.56), and 68 ml/min (AUC: 0.81, sensitivity=1.00, and specificity=0.68), respectively. We excluded BUN (COV:17 mg/dl, AUC: 0.74) because it had lower accuracy and was a confounding factor for eGFR. We performed univariate logistic analysis incorporating each factor: statin use and six laboratory factors. This analysis revealed that elevated LDH (OR: 14, P=0.0188), decreased eGFR (OR: 12.8, P=0.0233), decreased lymphocyte count (OR: 9.8, P=0.0414), and statin use (OR: 7.0, P=0.0230) were significantly associated with disease progression (Table 4).

Similarly, a plot of the progression-free intervals of the 57 patients is shown in Figure 2-A. Univariate Cox regression analysis showed that statin use (HR: 6.28, P=0.0167), elevated LDH (HR: 13.3, P=0.0169), and decreased eGFR (OR: 10.2 P=0.0315) but not lymphocytopenia were significantly associated with disease progression (Figure 2).

**Table 3.**
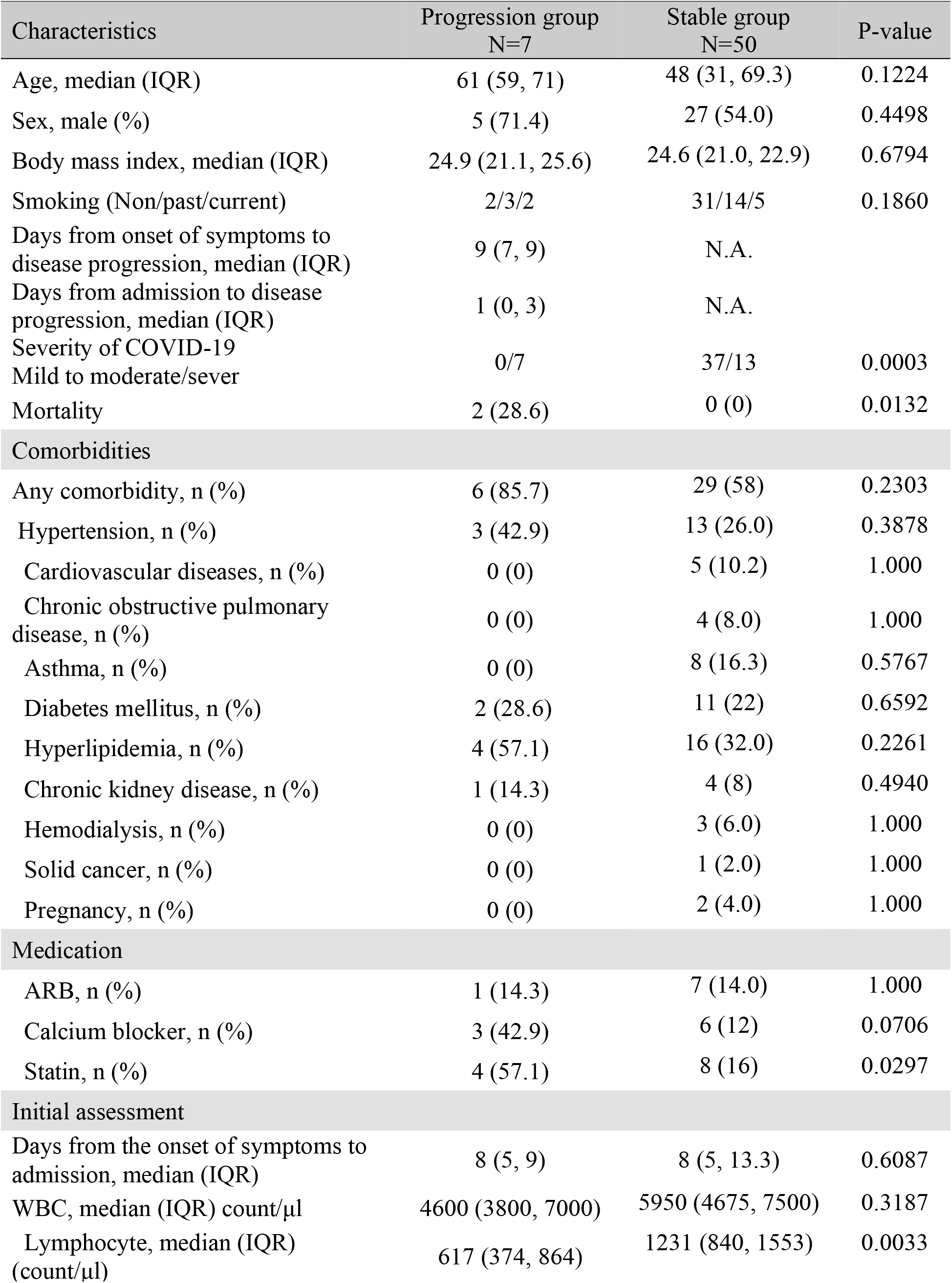
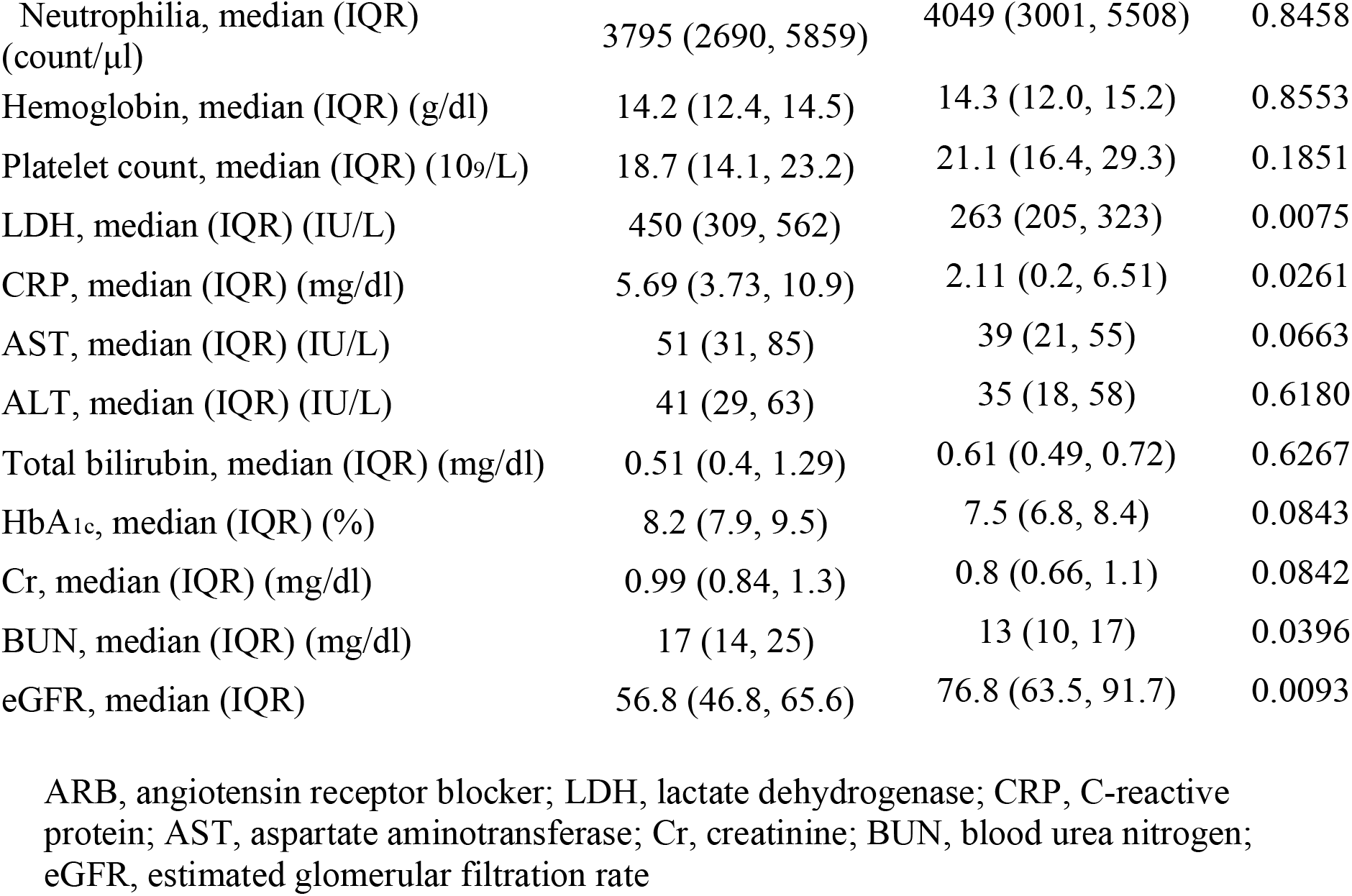
Comparison of the progression group and stable group.

| Characteristics | Progression group<br>N=7 | Stable group<br>N=50 | P-value |
| --- | --- | --- | --- |
| Age, median (IQR) | 61 (59, 71) | 48 (31, 69.3) | 0.1224 |
| Sex, male (%) | 5 (71.4) | 27 (54.0) | 0.4498 |
| Body mass index, median (IQR) | 24.9 (21.1, 25.6) | 24.6 (21.0, 22.9) | 0.6794 |
| Smoking (Non/past/current) | 2/3/2 | 31/14/5 | 0.1860 |
| Days from onset of symptoms to disease progression, median (IQR) | 9 (7, 9) | N.A. |  |
| Days from admission to disease progression, median (IQR) | 1 (0, 3) | N.A. |  |
| Severity of COVID-19<br>Mild to moderate/sever | 0/7 | 37/13 | 0.0003 |
| Mortality | 2 (28.6) | 0 (0) | 0.0132 |
| **Comorbidities** |  |  |  |
| Any comorbidity, n (%) | 6 (85.7) | 29 (58) | 0.2303 |
| Hypertension, n (%) | 3 (42.9) | 13 (26.0) | 0.3878 |
| Cardiovascular diseases, n (%) | 0 (0) | 5 (10.2) | 1.000 |
| Chronic obstructive pulmonary disease, n (%) | 0 (0) | 4 (8.0) | 1.000 |
| Asthma, n (%) | 0 (0) | 8 (16.3) | 0.5767 |
| Diabetes mellitus, n (%) | 2 (28.6) | 11 (22) | 0.6592 |
| Hyperlipidemia, n (%) | 4 (57.1) | 16 (32.0) | 0.2261 |
| Chronic kidney disease, n (%) | 1 (14.3) | 4 (8) | 0.4940 |
| Hemodialysis, n (%) | 0 (0) | 3 (6.0) | 1.000 |
| Solid cancer, n (%) | 0 (0) | 1 (2.0) | 1.000 |
| Pregnancy, n (%) | 0 (0) | 2 (4.0) | 1.000 |
| **Medication** |  |  |  |
| ARB, n (%) | 1 (14.3) | 7 (14.0) | 1.000 |
| Calcium blocker, n (%) | 3 (42.9) | 6 (12) | 0.0706 |
| Statin, n (%) | 4 (57.1) | 8 (16) | 0.0297 |
| **Initial assessment** |  |  |  |
| Days from the onset of symptoms to admission, median (IQR) | 8 (5, 9) | 8 (5, 13.3) | 0.6087 |
| WBC, median (IQR) count/μl | 4600 (3800, 7000) | 5950 (4675, 7500) | 0.3187 |
| Lymphocyte, median (IQR) (count/μl) | 617 (374, 864) | 1231 (840, 1553) | 0.0033 |
| Neutrophilia, median (IQR)<br>(count/μl) | 3795 (2690, 5859) | 4049 (3001, 5508) | 0.8458 |
| Hemoglobin, median (IQR) (g/dl) | 14.2 (12.4, 14.5) | 14.3 (12.0, 15.2) | 0.8553 |
| Platelet count, median (IQR) ($10^9$/L) | 18.7 (14.1, 23.2) | 21.1 (16.4, 29.3) | 0.1851 |
| LDH, median (IQR) (IU/L) | 450 (309, 562) | 263 (205, 323) | 0.0075 |
| CRP, median (IQR) (mg/dl) | 5.69 (3.73, 10.9) | 2.11 (0.2, 6.51) | 0.0261 |
| AST, median (IQR) (IU/L) | 51 (31, 85) | 39 (21, 55) | 0.0663 |
| ALT, median (IQR) (IU/L) | 41 (29, 63) | 35 (18, 58) | 0.6180 |
| Total bilirubin, median (IQR) (mg/dl) | 0.51 (0.4, 1.29) | 0.61 (0.49, 0.72) | 0.6267 |
| HbA<sub>1c</sub>, median (IQR) (%) | 8.2 (7.9, 9.5) | 7.5 (6.8, 8.4) | 0.0843 |
| Cr, median (IQR) (mg/dl) | 0.99 (0.84, 1.3) | 0.8 (0.66, 1.1) | 0.0842 |
| BUN, median (IQR) (mg/dl) | 17 (14, 25) | 13 (10, 17) | 0.0396 |
| eGFR, median (IQR) | 56.8 (46.8, 65.6) | 76.8 (63.5, 91.7) | 0.0093 |

ARB, angiotensin receptor blocker; LDH, lactate dehydrogenase; CRP, C-reactive protein; AST, aspartate aminotransferase; Cr, creatinine; BUN, blood urea nitrogen; eGFR, estimated glomerular filtration rate

**Table 4.**
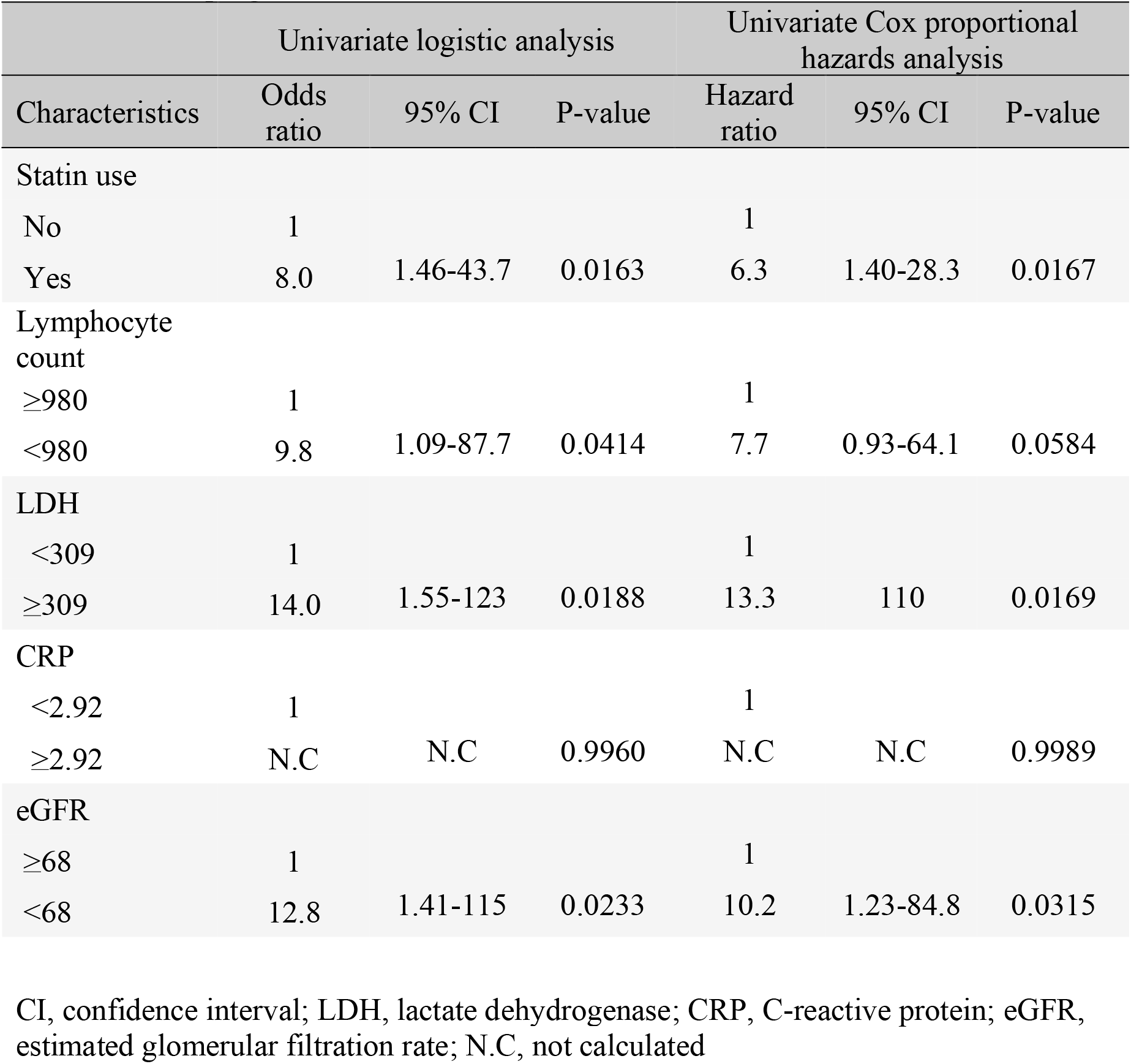
Univariate logistic analysis and univariate Cox proportional hazards analysis of risk factors for progression to critical COVID-19

**Figure 2.**
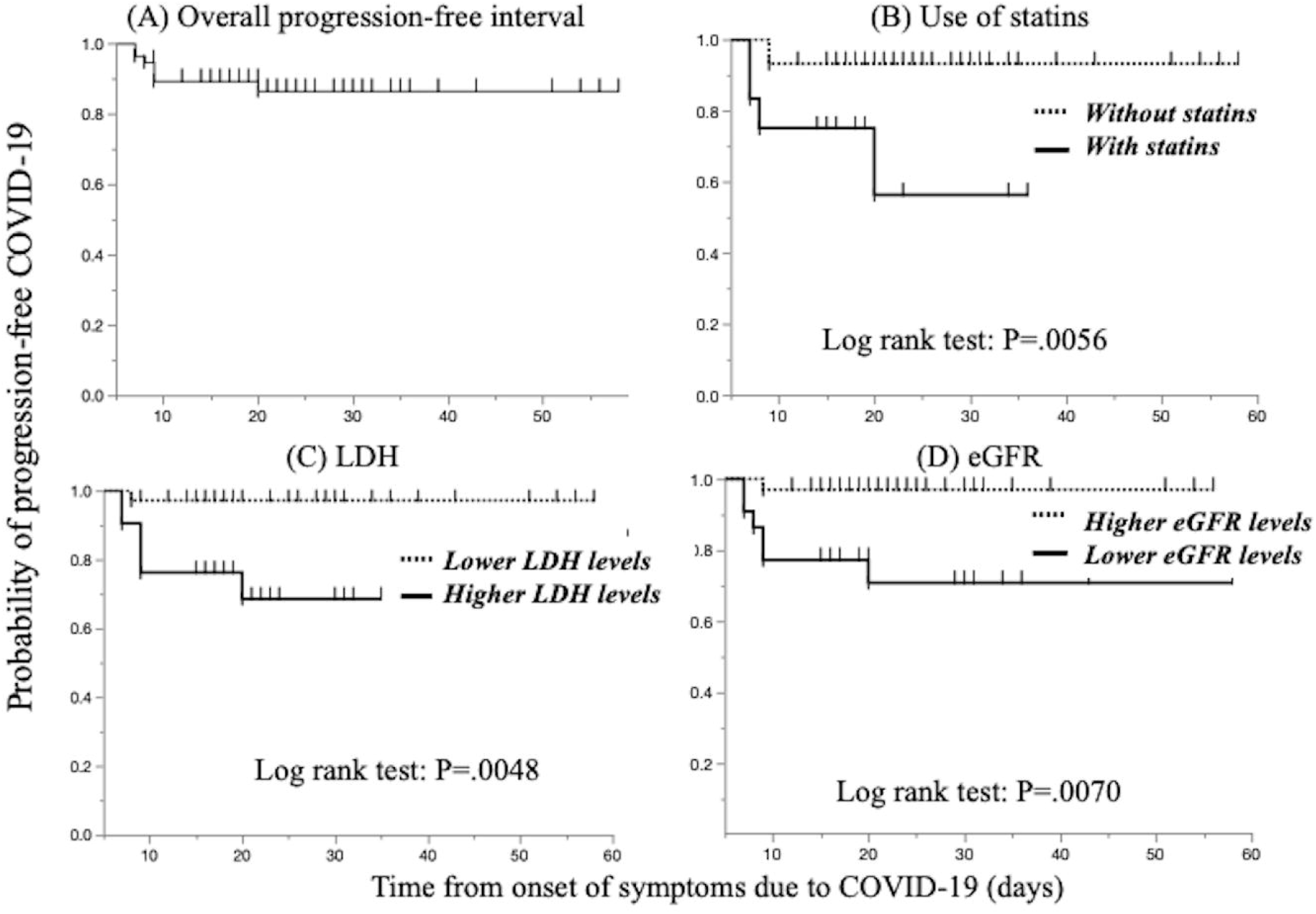
Overall progression-free interval in patients with mild to moderate COVID-19. Significant differences were found in the progression-free interval between patients stratified by statin use, LDH levels and eGFR on admission.

## Discussion

This study aimed to evaluate early predictive clinical factors for the progression of mild to moderate COVID-19 to critical COVID-19 in a Japanese patient population. To date, several laboratory findings have been reported to be associated with worse outcomes: lower lymphocyte counts, elevated liver enzymes, elevated LDH levels, high levels of inflammatory markers (e.g., CRP and ferritin), elevated D-dimer, prolonged prothrombin time, and acute kidney injury ^7,8^. Our results were consistent with those in previous reports. We identified four candidate risk factors for disease progression in patients with mild to moderate COVID-19—elevated LDH, statin use, decreased eGFR, and decreased lymphocyte count—by univariate logistic analysis, and three of these factors (excluding lymphocytopenia) significantly differed when time-to-event was considered. Unlike statin use, LDH and eGFR considered organ damage markers but not inflammatory factors. Recent studies have reported that severe COVID-19 is commonly complicated with coagulopathy and that disseminated intravascular coagulation may be involved in most deaths ^9^. An elevated LDH level and a decreased eGFR can indicate liver and kidney dysfunction. In addition, CRP, as an inflammatory marker, had higher sensitivity (100%) but lower specificity (56%) than other inflammatory markers. Therefore, we think that an elevated LDH level and a decreased eGFR are better essential risk factors predicting progression to critical disease or death than are inflammatory markers.

Regarding clinical factors, advanced age and male sex were reported to be significant risk factors for disease progression ^10-12^, but we did not identify these clinical factors as risk factors. Mortality risks have been reported as advanced age and comorbid conditions such as diabetes ^1,7,13,^ although severe disease can occur even in healthy individuals of any age ^13,14^. In the present study, we did not identify clinical background parameters as risk factors; however, both of our patients who died were men with diabetes aged 70 years or older, which supports the above previous reports.

In the present study, statin use was identified as a significant risk factor for developing critical disease. Statins have anti-inflammatory, antithrombotic, and immunomodulatory effects ^15,16^. Two observational studies of hospitalized patients with influenza during the 2009 H1N1 pandemic revealed reductions of 41% ^17^ and 59% ^18^ in 30-day mortality associated with the use of statins. Those studies showed that statin use could decrease the disease severity in hospitalized patients with influenza. However, a later investigation by Laidler et al. concluded that statins should not be used as an adjunct treatment to prevent death, because of unmeasured confounding ^18^. We agree with Laidler’s conclusion, because our cohort showed negative results with statin use, and our patients who used statins were of an advanced age (median, 62 years) and had comorbidities.

This report is the first to evaluate risk factors for disease progression in Japanese patients with COVID-19. Risk factors may be related to ethnic differences. In symptomatic Japanese cohort in this study, 65% had mild to moderate disease, and 35% had severe disease; the condition of 12.2% of these patients deteriorated, and two died (3.5%). In a large cohort of Chinese patients, including approximately 44,500 patients with confirmed COVID-19, 81% of cases were classified as mild, 14% as severe, and 5% as critical, and the mortality rate was 2.3% ^13^. The outcomes in our Japanese cohort were similar to or worse than those in this Chinese population, and there seemed to be no ethnic differences, considering the triaged patients in our cohort.

This study has several limitations due to its retrospective nature. First, we enrolled only a small number of COVID-19 patients. Consequently, we did not perform multivariate analysis. Second, our dataset had missing data, because we were not very accustomed to seeing COVID-19 patients and avoided unnecessary or nonurgent contact with these patients to reduce the risk of infection. Therefore, we statistically handled missing data using imputation methods in statistical software.

In conclusion, we first reported three candidate risk factors in Japanese adult patients with mild to moderate COVID-19: statin use, an elevated LDH level, and decreased eGFR.

## Data Availability

Data is available upon request.

## Acknowledgments and collaborators

We thank all medical staff and doctors at Toyonaka Municipal Hospital. The collaborators involved in the study are as follows: Sanae Fukuda, Kazumi Ohkubo (Nursing Department), Dr. Masashi Yamamoto, Dr. Kengo Matsumoto, Dr. Kaori Mukai, Dr. Dai Nakamatsu, Dr. Aya Sugimoto, Dr. Naoto Osugi, Dr. Sho Yamaoka, Dr. Tatsuya Sakamoto, Dr. Akino Okamoto, Dr. Yuri Tsujii, Dr. Ryo Sugio, Dr. Kazumasa Souma (Department of Gastroenterology), Dr. Masayuki Moriya, Dr. Katsuya Araki, Dr. Yuri Sugiura (Department of Neurology), Dr. Masanobu Takeji, Dr. Satoko Yamamoto, Dr. Yasuo Kusunoki, Dr. Natsuko Ikeda, Dr. Kumie Teramoto, Dr. Momoko Okawara, Dr. Yuki Iwahashi, Dr. Masashi Yokoyama, Dr. Toru Kida, Dr. Chihiro Hasegawa, Dr. Shunsuke Shiode, Dr. Tomoko Isaka, Dr. Naohiko Ito, Dr. Kanae Matsuno (Department of Internal Medicine), Dr. Yukinori Okazaki, Dr. Yukika Mizukami, Dr. Takuma Iida, Dr. Naoki Fukushima, Dr. Ai Miyaoka, Dr. Takamori Yamamoto (Department of Cardiology).

